# Plasma SARS-CoV-2 RNA levels as a biomarker of lower respiratory tract SARS-CoV-2 infection in critically ill patients with COVID-19

**DOI:** 10.1101/2022.01.10.22269018

**Authors:** Jana L. Jacobs, Asma Naqvi, Faraaz A. Shah, Valerie F. Boltz, Mary F. Kearney, Bryan J. McVerry, Prabir Ray, Caitlin Schaefer, Meghan Fitzpatrick, Barbara Methé, Janet Lee, Alison Morris, John W. Mellors, Georgios D. Kitsios, William Bain

**Author notes:** Address correspondence to: Jana L Jacobs, Division of Infectious Diseases, Department of Medicine, University of Pittsburgh School of Medicine, address 3550 Terrace St, Ste 814 Scaife Hall, Pittsburgh, PA 15261 USA. these authors contributed equally (Georgios D. Kitsios, William Bain).

## Abstract

Plasma SARS-CoV-2 viral RNA (vRNA) levels are predictive of COVID-19 outcomes in hospitalized patients, but whether plasma vRNA reflects lower respiratory tract (LRT) vRNA levels is unclear. We compared plasma and LRT vRNA levels in simultaneously collected longitudinal samples from mechanically-ventilated patients with COVID-19. LRT and plasma vRNA levels were strongly correlated at first sampling (r=0.83, p<10^−8^) and then declined in parallel except in non-survivors who exhibited delayed vRNA clearance in LRT samples. Plasma vRNA measurement may offer a practical surrogate of LRT vRNA burden in critically ill patients, especially early in severe disease.

## Background

Plasma SARS-CoV-2 RNA (vRNA) levels (RNAemia) are significantly associated with COVID-19 severity indices and predict adverse clinical outcomes in hospitalized patients [1-6]. We and others have recently shown that detectable RNAemia can indicate presence of virions in plasma (i.e. true viremia) [1, 7], but how RNAemia relates to vRNA levels in the lower respiratory tract (LRT) has not been well defined. LRT infection by SARS-CoV-2 can lead to pneumonia and acute respiratory distress syndrome (ARDS), which is the most common cause of COVID-19 mortality [8]. Viral particles are readily detectable by electron microscopy in mechanically ventilated patients with COVID-19, and vRNA levels in LRT secretions have been shown to be associated with COVID-19 outcome [9-11], but the relationship between LRT and plasma vRNA is not well defined. Levels of plasma vRNA could be an important biomarker of the extent of LRT infection. To explore this possibility, we compared plasma and LRT vRNA levels in simultaneously-collected specimens, and examined their associations and relations to clinical outcomes.

## Methods

### Study Cohort

From April 2020 through May 2021, we prospectively enrolled hospitalized patients with COVID-19 from three UPMC hospitals, in an observational cohort study. We included critically ill patients 18-90 years of age diagnosed with SARS-CoV-2 infection by a positive nasopharyngeal swab quantitative polymerase chain reaction (qPCR) test, who were intubated and mechanically ventilated for acute hypoxemic respiratory failure due to COVID-19 pneumonia.

#### IRB Approvals

All research protocols (protocols STUDY19050099 and STUDY20040036) were approved by University of Pittsburgh institutional review board and were performed in accordance with the Declaration of Helsinki. Written informed consent was obtained from all research participants or their legally authorized representatives.

### Clinical Data Extraction

We recorded baseline demographics, COVID-19 timelines (dates of symptom onset, SARS-CoV-2 infection diagnosis, ICU admission and intubation), severity indices at time of ICU admission (WHO 10-point ordinal scale and radiographic edema by the Radiographic Assessment of Lung Edema [RALE] score [12]), administered COVID-19-targeted therapies with plausible impact on vRNA levels (remdesivir, convalescent plasma, corticosteroids and tocilizumab), 60-day survival and time to liberation from mechanical ventilation.

### Experimental Analyses

We simultaneously collected serial blood samples and endotracheal aspirates (ETA) on enrollment day (Day 1 - baseline), and then again on Days 5 and 10 post-enrollment while the subjects remained hospitalized in the ICU. Blood samples were centrifuged for separation and storage of plasma. ETA samples were inactivated and RNA was preserved by mixing at 1:3 volume ratio with Zymo DNA/RNA shield under Biosafety Level 2+ conditions and then stored at -80°C until testing.

#### Quantitative RT-PCR for SARS-CoV-2

We performed qRT-PCR for SARS-CoV-2 as previously described for plasma samples [1]. Briefly, we extracted total RNA from 0.5-1.0 mL of plasma using the MagMax Viral Pathogens Kit and the Kingfisher Flex automated extractor (Thermofisher). For ETA samples, we diluted 150µl of the inactivated ETA sample with 250µl phosphate buffered saline to final volume of 400µL to reduce ETA viscosity, and then extracted total RNA using the MagMax Viral Pathogens Kit and the Kingfisher Flex automated extractor (Thermofisher). We performed subsequent one-step quantitative RT-PCR of SARS-CoV-2 N and human RNaseP as previously described using the following N-specific primers (Fwd: 5’-GTTTGGTGGACCCTCAGATT-3’, Rev: 5’-CGCAGTATTATTGGGTAAACCTTG-3’, Probe: 5’6-FAM-TAACCAGAATGGAGAACGCAGTGGG-3’BHQ1) [1].

### Statistical Analyses

We performed log_10_-transformed vRNA level comparisons between sample types and clinical groups with non-parametric Wilcoxon tests. We examined plasma-ETA vRNA correlations with the Spearman rank method. We examined the dynamics of vRNA levels over time in linear regression models of plasma or ETA vRNA against days of sample acquisition from symptom onset, PCR diagnosis, ICU admission, and intubation. For the time-to-event outcomes of 60-day survival and time to liberation from IMV, we constructed Cox proportional hazards models with plasma or ETA vRNA as exposure, adjusted for age and time from symptom onset. We limited clinical outcome analyses to include only those samples collected within six days from ICU admission to account for immortal time bias. We analyzed temporal evolution of vRNA levels in mixed linear regression models with random patient intercepts.

## Results

### Comparisons of plasma and ETA vRNA levels

We included 54 subjects (median age 63 years, 63% men), who contributed a total of 96 plasma and 77 ETA samples (Fig S1). Clinical characteristics in 60-day survivors (n=28) vs. non-survivors (n=26) are shown in Table S1. We examined the first available sample pair (ETA and plasma) post-intubation among 54 patients treated with invasive mechanical ventilation (IMV) (analysis A, Fig S1). vRNA was detectable in 30/52 (57.7%) plasma and 38/47 (80.9%) ETA samples (p-value for difference in proportions = 0.01). The nine samples with undetectable ETA vRNA also had undetectable plasma vRNA (Fig 1A). Conversely, subjects with undetectable plasma vRNA had significantly lower ETA vRNA levels compared to subjects with detectable plasma vRNA (p<0.0001, Fig 1B).

**Figure 1.**
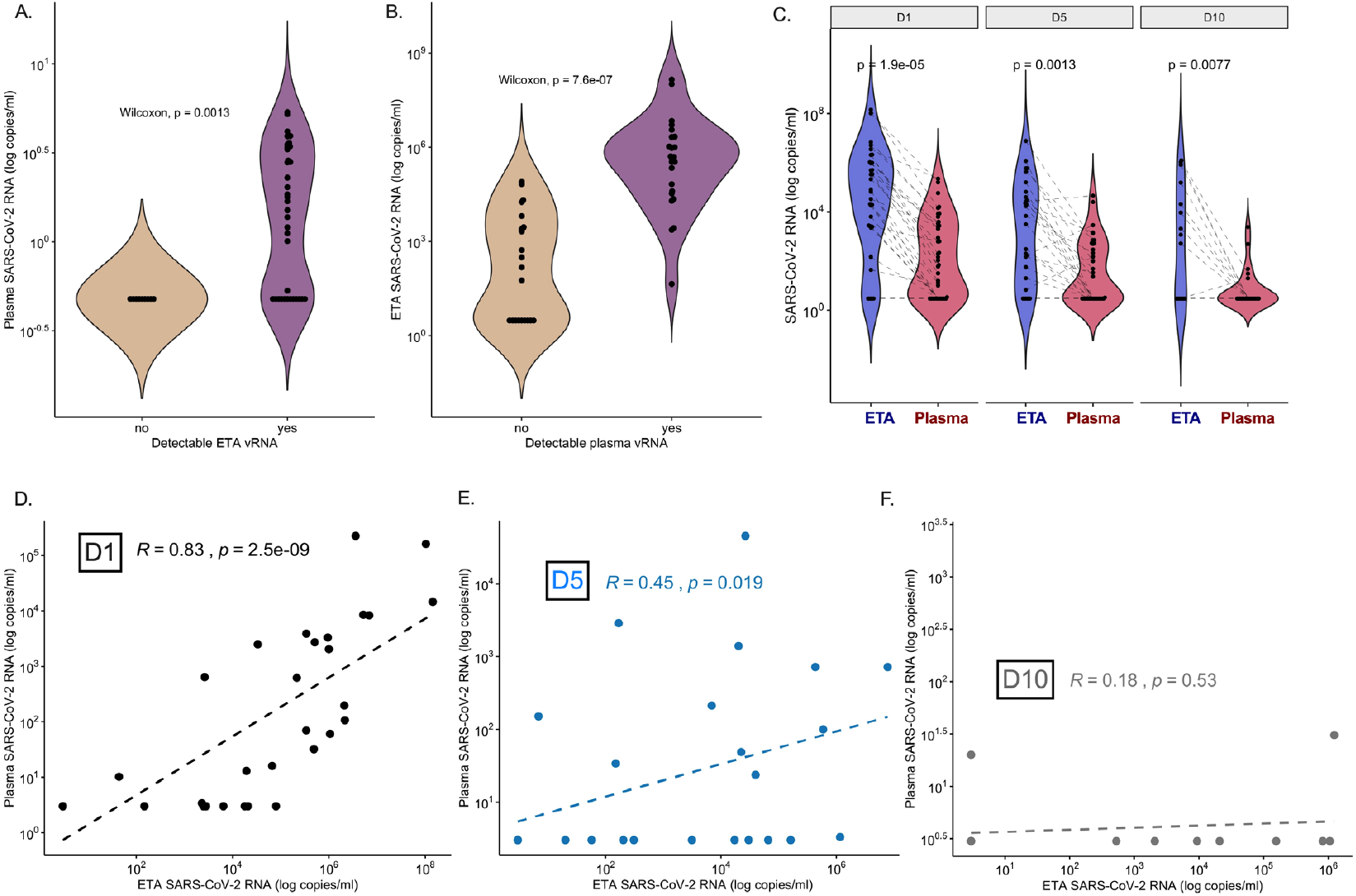
Plasma SARS-CoV-2 viral RNA levels reflect lower respiratory tract viral RNA levels. (A) Plasma vRNA levels in first available sample pair separated by non-detectable (left) or detectable (right) ETA vRNA. (B) ETA vRNA levels in first available sample pair separated by non-detectable (left) or detectable (right) plasma vRNA. Comparisons in (A) and (B) were done using Wilcoxon non-parametric test. (C) ETA and plasma vRNA levels by day of sampling (days 1, 5 and 10 post-enrollment). Each point represents a single sample and the dotted line indicates paired ETA and plasma samples. Comparisons in (A-C) by Wilcoxon test. (D-F): Scatter plots of paired ETA and plasma SARS-CoV-2 RNA levels stratified by sampling day (D: D1; E: D5, F: D10) with displayed linear regression lines and Spearman rank test correlations and p-values. Abbreviations: D1 = enrollment day 1. D5 = post-enrollment day 5. D1 = post-enrollment day 10. ETA: endotracheal aspirate. mL = milliliter. RNA = ribonucleic acid. SARS-CoV-2 = Severe acute respiratory syndrome coronavirus 2. vRNA = SARS-CoV-2 RNA.

Stratified by enrollment day (analysis B in Fig S1), ETA samples had significantly higher median number of vRNA copies compared to matched plasma samples per mL of specimen analyzed (D1 ratio of LRT to plasma vRNA: 635, D5: 31, D10: 175; all p<0.01, Fig 1C). In D1 sample pairs, ETA and plasma vRNA were strongly correlated (Spearman r=0.83, p<10^−8^), with attenuated correlation in D5 (r=0.45, p=0.02), and non-significant correlation by D10, when most plasma samples had undetectable vRNA (Fig. 1D-F).

### Plasma and lower respiratory tract vRNA by time from symptom onset

We then examined for the impact of the recorded COVID-19 timeline on plasma and ETA vRNA levels in mixed linear regression models of vRNA against time (days) from symptom onset for all samples available. Both ETA and plasma vRNA levels significantly decreased over time (adjusted p<0.001, Fig 2A), with all plasma samples becoming undetectable for vRNA by post-symptom day 23. We observed similar significant decrements of vRNA levels over time when considering time periods from SARS-CoV-2 diagnosis, ICU admission and intubation (data not shown).

**Figure 2.**
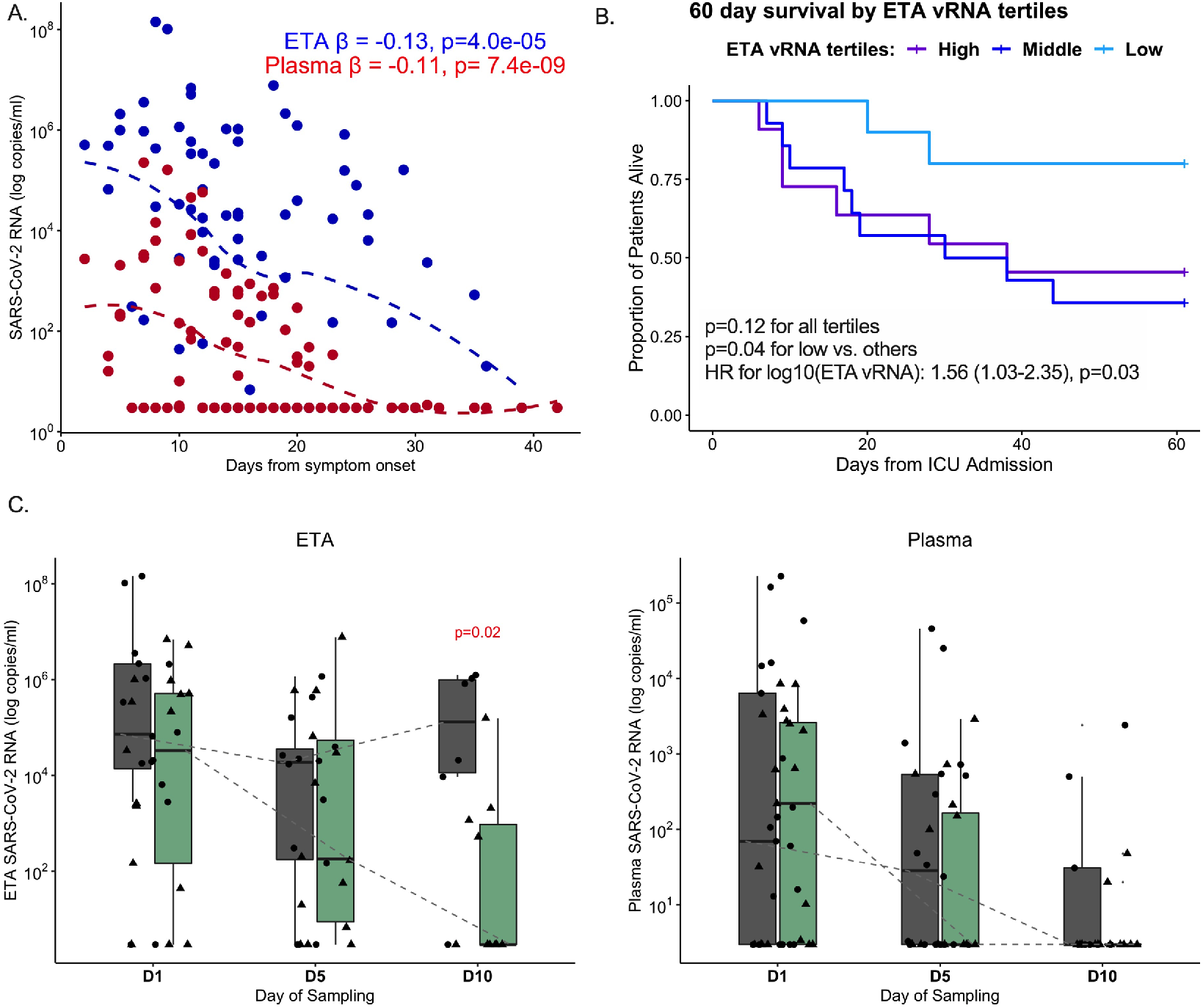
Plasma and lower respiratory tract SARS-CoV-2 viral levels exhibit similar temporal decline although persistently elevated RNA levels in lower respiratory tract samples are associated with increased mortality. (A) Scatterplot of reported days from COVID-19 symptom onset (x-axis) and viral RNA (vRNA) levels (log_10_-transformed) in endotracheal aspirate (blue) and plasma (red) samples. The beta coefficients (β) and corresponding p-values of mixed linear regression models of log_10_-transformed vRNA levels with random patient intercepts and adjustment for time of sample acquisition from symptom onset are shown, with displayed dashed lines from locally weighted scatterplot smoothing. (B) Kaplan-Meier curves of 60-day survival by ETA vRNA tertiles among patient samples obtained within the first six days of ICU admission: low tertile<4,844; middle 4,482-490,922; high> 490,922 copies/ml. (C and D) ETA and plasma vRNA levels, stratified by sampling day and comparing non-survivors (dark gray) and survivors (light green). Statistical comparisons for differences in trajectories were performed with mixed linear regression models with random patient intercepts and interaction terms for survivorship*sampling day. Abbreviations: D1 = enrollment day 1. D5 = post-enrollment day 5. D1 = post-enrollment day 10. ETA: endotracheal aspirate. HR = hazard ratio. mL = milliliter. RNA = ribonucleic acid. SARS-CoV-2 = Severe acute respiratory syndrome coronavirus 2. vRNA = SARS-CoV-2 RNA.

### Associations of vRNA with Outcomes

By examining only samples obtained within six days of ICU admission (analysis C in Fig S1, n=47 plasma and n=35 ETA unique subject samples), we found no significant association of either plasma or ETA vRNA levels with cross-sectional indices of COVID-19 severity (WHO ordinal scale or radiographic edema scores). Furthermore, we found no significant association for either plasma or ETA vRNA levels with receipt of the examined COVID-19 therapies prior to sample acquisition (data not shown).

ETA vRNA levels among samples obtained within six days of ICU admission were significantly associated with 60-day survival in a Cox proportional hazards model adjusted for age and time (days) of sample acquisition from symptom onset (hazard ratio [HR], 95% confidence interval = 1.56 [1.03-2.35] for log_10_-transformed ETA vRNA levels, p=0.03). By stratifying early ETA vRNA levels (within six days of ICU admission) in tertiles and examining 60-day survival in Kaplan-Meier curve analysis, we found that patients in the low tertile (below 4,842 copies/ml) had improved survival compared to patients to the middle (between 4,482-490,922 copies/ml) and high (above 490,922 copies/ml) tertiles combined (log-rank p=0.04, Fig 2B). Similarly, higher early ETA vRNA levels were predictive of longer times for liberation from mechanical ventilation among 60-day survivors in an adjusted Cox proportional hazards model (HR for successful liberation per log10-transformed vRNA levels= 0.81 [0.65-1.00], p=0.05). Early plasma vRNA levels were not significantly associated with worse 60-day survival or liberation from mechanical ventilation, although HR point estimates were consistent in direction with the effects observed for ETA vRNA levels (1.18 and 0.82, respectively).

We then examined the vRNA level trajectories of available longitudinal samples between 60-day survivors and non-survivors in mixed linear regression models with interaction terms for 60-day mortality and day of sampling (Fig 2C-D). We detected no significant difference in vRNA level decline for survivors and non-survivors (interaction term p-values 0.06 and 0.43 for ETA and plasma samples respectively). However, non-survivors had significantly higher ETA vRNA levels in D10 samples (p=0.02, Fig 2C), signifying delayed vRNA clearance compared to survivors. There was no difference in D10 plasma samples between survivors and non-survivors, as by D10 17/22 (77.8%) of samples had become undetectable for vRNA.

## Discussion

We show that SARS-CoV-2 viral RNA levels in plasma and LRT secretions are strongly correlated in patients with severe COVID-19 early after ICU admission. This finding provides support for plasma vRNA as an indicator of lung SARS-CoV-2 infection and suggests that plasma vRNA may be a useful biomarker for LRT viral burden. A practical blood marker of LRT viral infection is desirable to improve sample accessibility and increase standardization of inter-patient collection methodology because there may be greater potential variability in LRT secretion collection.

We did not demonstrate a difference in clinical outcomes based on plasma vRNA even though we and several others have previously published that plasma vRNA is associated with, and predictive of, clinical outcomes [1-6]. This divergent result is likely due to analyses restricted only to critically ill COVID-19 patients in the ICU. Prior analyses showing associations of plasma vRNA with clinical outcome included subjects across a much wider spectrum of COVID-19 severity and patients earlier in their disease course [1-6]. We noted a wide range in the time interval between symptom onset and admission to the ICU in the current study (median=7, range of 2-31 days), indicating highly variable COVID-19 disease course. We show that by the time patients were admitted to the ICU and enrolled in our study, a higher proportion of plasma samples had become undetectable for vRNA compared to ETA samples (42.3% vs. 19.1%, p=0.01). We speculate that a larger sample size of plasma could have revealed a significant predictive effect given the proportionally similar increase in plasma vRNA when comparing non-survivors to survivors

Consistent with prior evidence [13, 14], we showed that both LRT and plasma vRNA decrease over time, with higher levels and prolonged detection of LRT vRNA in non-survivors compared to survivors. Furthermore, we show statistically significant parallel declines in LRT and plasma vRNA levels from the time of symptom onset, which supports the feasibility of plasma vRNA as a biomarker of LRT vRNA in critically ill patients. Our findings support the importance of adjusting for time from symptom onset in survival analyses, given the variability in timings of patient presentation and study enrollment during the clinical course of COVID-19.

The biological mechanisms that underlie the strong correlation between vRNA levels in the LRT and blood compartment are unclear. One potential pathway by which virions transit from lung to blood is disruption of the air-blood barrier due to inflammation and/or direct viral injury allowing spillover of virions from lung to bloodstream [15]. Regardless of the mechanism, transit of virions from the LRT to the bloodstream may lead to extrapulmonary dissemination of infection; indeed others have shown presence of extrapulmonary infectious virus [7].

Our analyses are limited by cohort size and inclusion of only ICU patients. Nevertheless, the well-defined cohort of participants we studied allowed for direct, minimally-invasive access to the LRT for vRNA quantification in ETA specimens, which are routinely obtained for clinical microbiology studies in COVID-19 subjects, as opposed to invasive bronchoscopic samples. We did not analyze upper respiratory tract vRNA levels, because our focus was to examine LRT viral burden and its relationship with plasma vRNA levels in patients with severe disease, and viral shedding in severe COVID-19 has been reported to be prolonged in the lower as compared to the upper respiratory tract [11]. Our findings also highlight the importance of analyzing the timing of measurement of vRNA biomarkers in relation to the clinical timeline of COVID-19, given the rapid and dynamic decline of vRNA levels in both the LRT and blood compartments.

In summary, plasma SARS-CoV-2 vRNA may serve as a useful biomarker of LRT infection in critically ill patients. Further research is necessary to confirm our findings in additional cohorts, using larger datasets to determine whether persistence of plasma viremia in critically ill patients is associated with worse clinical outcomes, and to investigate the biological mechanisms that underlie the relationship between lung and plasma viral burden.

## Data Availability

All data produced in the present study are available upon reasonable request to the authors

## Acknowledgments

The authors wish to thank the patients and patient families that have enrolled in our research studies at the University of Pittsburgh. We also thank the physicians, nurses, respiratory therapists and other staff at the UPMC Presbyterian, Shadyside and East Hospital units for assistance with coordination of patient enrollment and collection of patient samples. We thank Heather Michael, Cathy Kessinger and Cynthia Klamar for assistance with patient enrollment and processing research samples.

## Supplemental Data

**Table S1:**
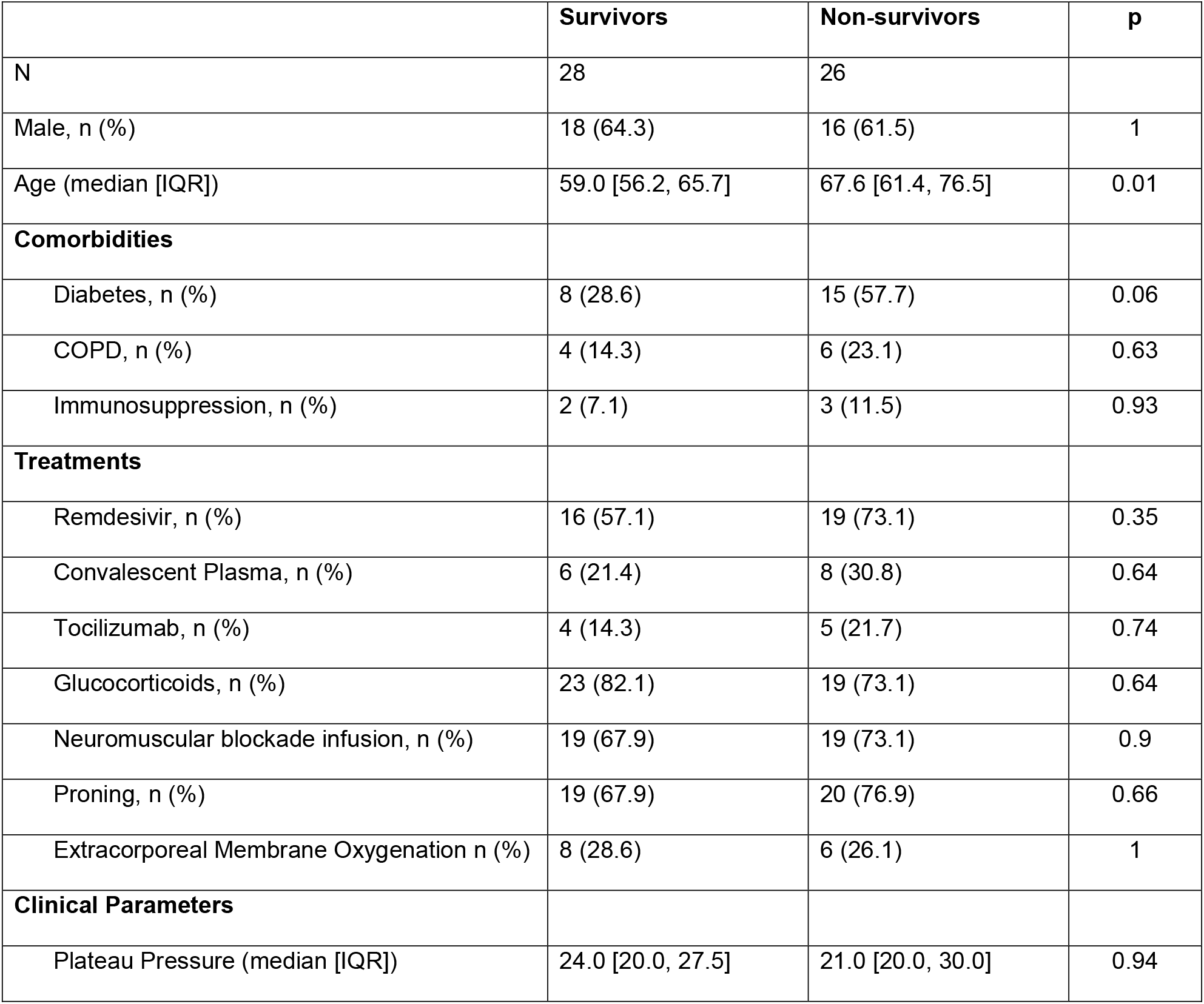
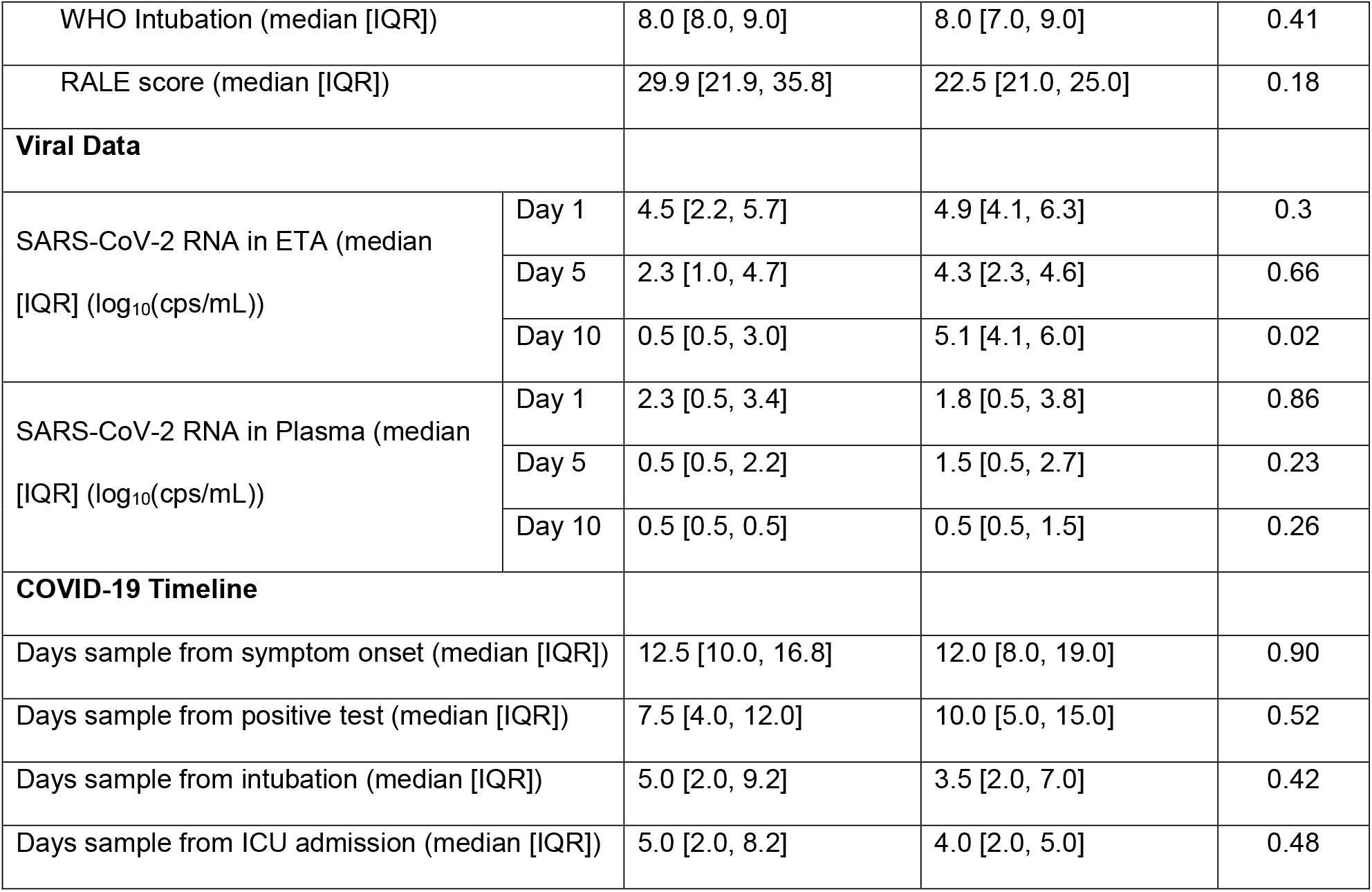
Clinical characteristics stratified by survival status at 60-days post ICU admission. P-values are derived from non-parametric tests (Wilcoxon tests for continuous and Fisher’s exact tests for categorical variables). Abbreviations: COPD = chronic obstructive pulmonary disease. IQR = inter-quartile range. RALE = radiographic assessment of lung edema. WHO = World Health Organization.

|  | **Survivors** | **Non-survivors** | **p** |
| --- | --- | --- | --- |
| N | 28 | 26 |  |
| Male, n (%) | 18 (64.3) | 16 (61.5) | 1 |
| Age (median [IQR]) | 59.0 [56.2, 65.7] | 67.6 [61.4, 76.5] | 0.01 |
| **Comorbidities** |  |  |  |
| Diabetes, n (%) | 8 (28.6) | 15 (57.7) | 0.06 |
| COPD, n (%) | 4 (14.3) | 6 (23.1) | 0.63 |
| Immunosuppression, n (%) | 2 (7.1) | 3 (11.5) | 0.93 |
| **Treatments** |  |  |  |
| Remdesivir, n (%) | 16 (57.1) | 19 (73.1) | 0.35 |
| Convalescent Plasma, n (%) | 6 (21.4) | 8 (30.8) | 0.64 |
| Tocilizumab, n (%) | 4 (14.3) | 5 (21.7) | 0.74 |
| Glucocorticoids, n (%) | 23 (82.1) | 19 (73.1) | 0.64 |
| Neuromuscular blockade infusion, n (%) | 19 (67.9) | 19 (73.1) | 0.9 |
| Proning, n (%) | 19 (67.9) | 20 (76.9) | 0.66 |
| Extracorporeal Membrane Oxygenation n (%) | 8 (28.6) | 6 (26.1) | 1 |
| **Clinical Parameters** |  |  |  |
| Plateau Pressure (median [IQR]) | 24.0 [20.0, 27.5] | 21.0 [20.0, 30.0] | 0.94 |

|  |  |  |  |  |
| --- | --- | --- | --- | --- |
| WHO Intubation (median [IQR]) |  | 8.0 [8.0, 9.0] | 8.0 [7.0, 9.0] | 0.41 |
| RALE score (median [IQR]) |  | 29.9 [21.9, 35.8] | 22.5 [21.0, 25.0] | 0.18 |
| **Viral Data** |  |  |  |  |
| SARS-CoV-2 RNA in ETA (median [IQR] ($\log_{10}(\text{cps/mL})$)) | Day 1 | 4.5 [2.2, 5.7] | 4.9 [4.1, 6.3] | 0.3 |
|  | Day 5 | 2.3 [1.0, 4.7] | 4.3 [2.3, 4.6] | 0.66 |
|  | Day 10 | 0.5 [0.5, 3.0] | 5.1 [4.1, 6.0] | 0.02 |
| SARS-CoV-2 RNA in Plasma (median [IQR] ($\log_{10}(\text{cps/mL})$)) | Day 1 | 2.3 [0.5, 3.4] | 1.8 [0.5, 3.8] | 0.86 |
|  | Day 5 | 0.5 [0.5, 2.2] | 1.5 [0.5, 2.7] | 0.23 |
|  | Day 10 | 0.5 [0.5, 0.5] | 0.5 [0.5, 1.5] | 0.26 |
| **COVID-19 Timeline** |  |  |  |  |
| Days sample from symptom onset (median [IQR]) |  | 12.5 [10.0, 16.8] | 12.0 [8.0, 19.0] | 0.90 |
| Days sample from positive test (median [IQR]) |  | 7.5 [4.0, 12.0] | 10.0 [5.0, 15.0] | 0.52 |
| Days sample from intubation (median [IQR]) |  | 5.0 [2.0, 9.2] | 3.5 [2.0, 7.0] | 0.42 |
| Days sample from ICU admission (median [IQR]) |  | 5.0 [2.0, 8.2] | 4.0 [2.0, 5.0] | 0.48 |

**Fig S1:**
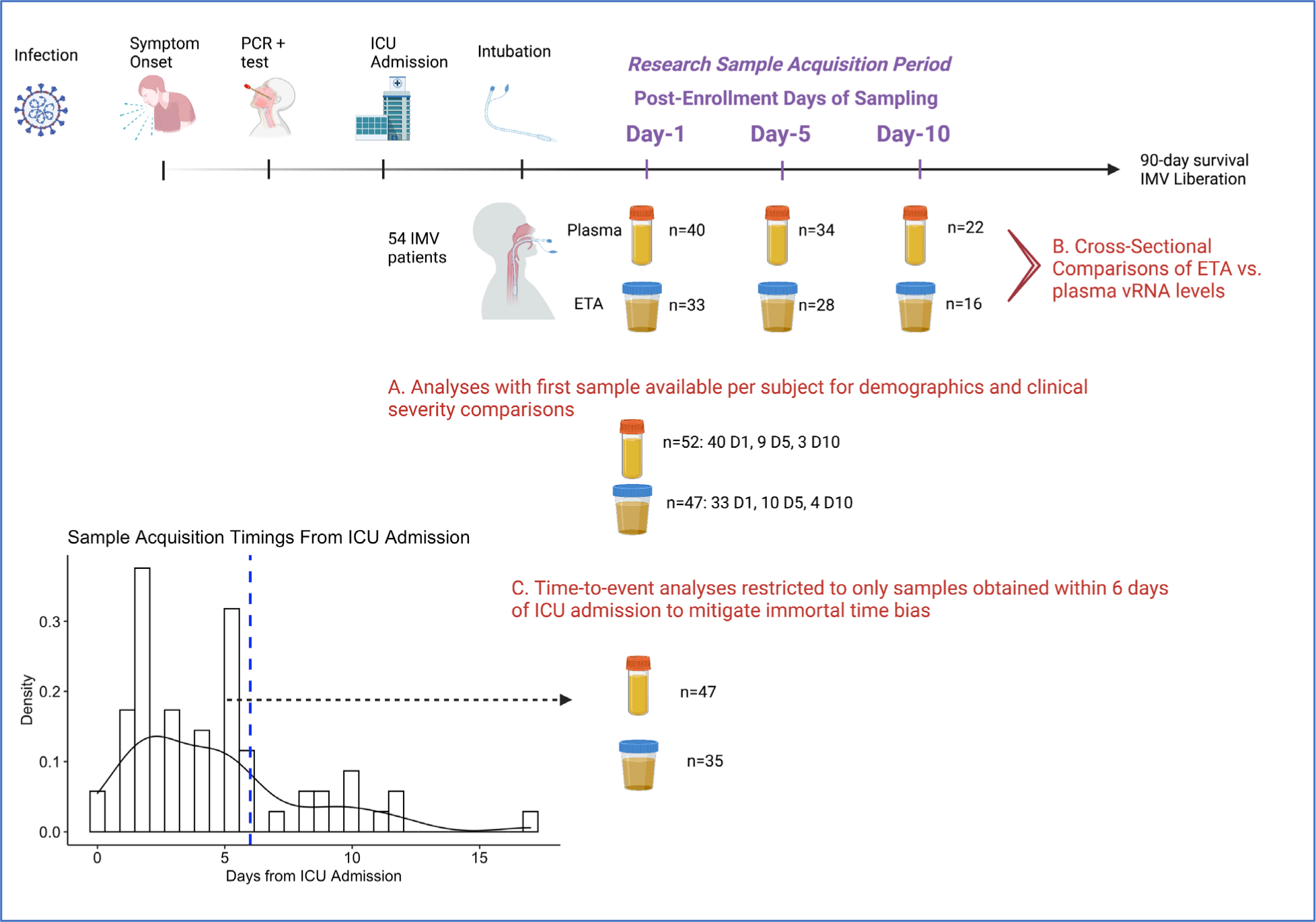
Graphical display of COVID-19 timeline, study samples, and key analyses.

